# The life course shaping of quality of life in older age: does childhood poverty associate with control, autonomy, self-realisation and pleasure? A retrospective and prospective study of HRS, ELSA, SHARE and TILDA cohorts

**DOI:** 10.64898/2026.11.04.26367287

**Authors:** Gindo Tampubolon

## Abstract

Later-life quality of life is increasingly recognised as a life-course achievement, yet ageing research has focused more on dysfunction and decline than on positive wellbeing. This study examines whether childhood poverty shapes later-life quality of life, measured by CASP: control, autonomy, self-realisation and pleasure.

**METHODS:** I analysed harmonised data from 59,569 adults aged over 50 in 29 countries, drawn from HRS, ELSA, SHARE and TILDA (U.S. Health and Retirement Study, English Longitudinal Study of Ageing, Survey of Health, Ageing and Retirement in Europe, The Irish Longitudinal Study on Ageing). Childhood poverty was modelled as a latent class from retrospectively reported childhood material circumstances, acknowledging recall error in such reports. Country fixed-effects linear models estimated associations with CASP, first adjusting for age, sex and country, then adding education, marital status and later-life wealth.

**RESULTS:** One in five participants grew up in poverty (22.9%). Mean CASP was 38 out of 48. Initially childhood poverty was associated with a 1.2-point lower CASP score; after complete adjustment the penalty attenuated but remained durable at 0.7 points. Education, partnership and wealth were independently associated with higher wellbeing but did not erase the penalty. Predicted age profiles showed poverty penalty in every country. They also revealed striking national differences: Nordic countries achieved markedly higher CASP levels than several Eastern European countries; Finns aged 90 had similar levels of positive wellbeing to Latvians two decades younger.

**CONCLUSION:** Childhood poverty leaves a durable mark on later-life positive wellbeing, while welfare contexts appear to lift overall CASP levels. Reducing child poverty and strengthening social protection are therefore lifelong wellbeing policies.

## INTRODUCTION

The twenty-first century is marked by demographic transformation: population is ageing rapidly around the world as profound social progress and medical advances continue.(1,2) This transformation poses wide-ranging challenges for states, societies and healthcare systems. What unfolded over a century in parts of Western Europe is now occurring within a few decades in Asia and beyond.(3) The geographic scope heightens the urgency of understanding not only how long people live, but how well they live. Global policy initiatives such as the World Health Organization’s (WHO) Decade of Healthy Ageing (2021–2030) have emphasised a shift away from disease treatment to the promotion of functional ability and wellbeing across the life-course.(4,5) This shift reflects a growing consensus that health in later-life cannot be understood other than as the culmination of cumulative exposures, experiences and transitions extending from pre-natal development through childhood, adulthood and into later-life.(6,7) The life-course framework has emerged as an ally to understand health inequalities in ageing societies.(8,9) Critical and sensitive periods, especially childhood, represent windows during which exposures can exert enduring effects on physiological systems and behavioural routines.(10) I have collected two frameworks on the life-course shaping of older adults’ (over 50 years) health (Figure 1, first, dementia from the Lancet Commission and second, broader outcomes from the WHO).(2) In comparison, the WHO framework appears more elaborate at the childhood stage and is used here to guide research highlighting the role of poor housing quality and overcrowding.

**Figure 1.**
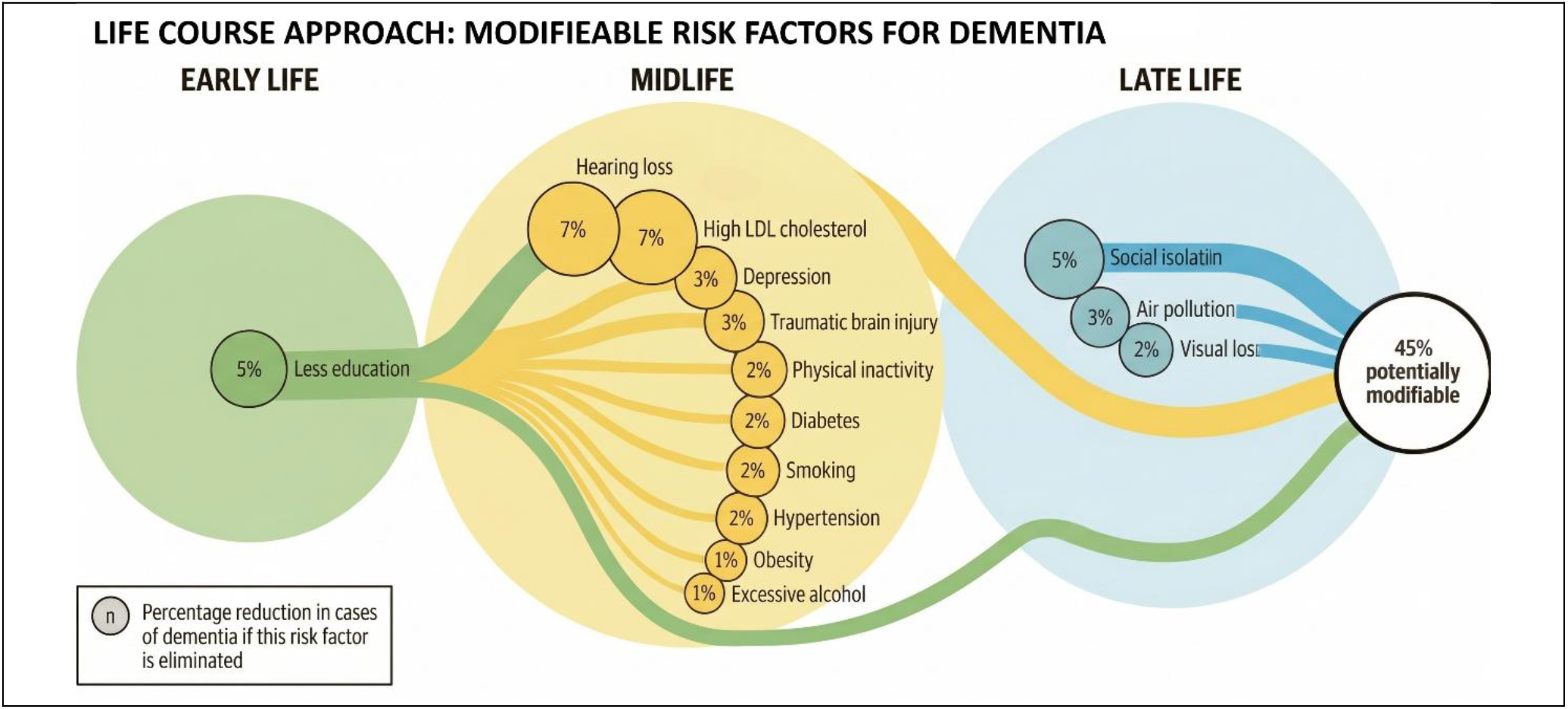
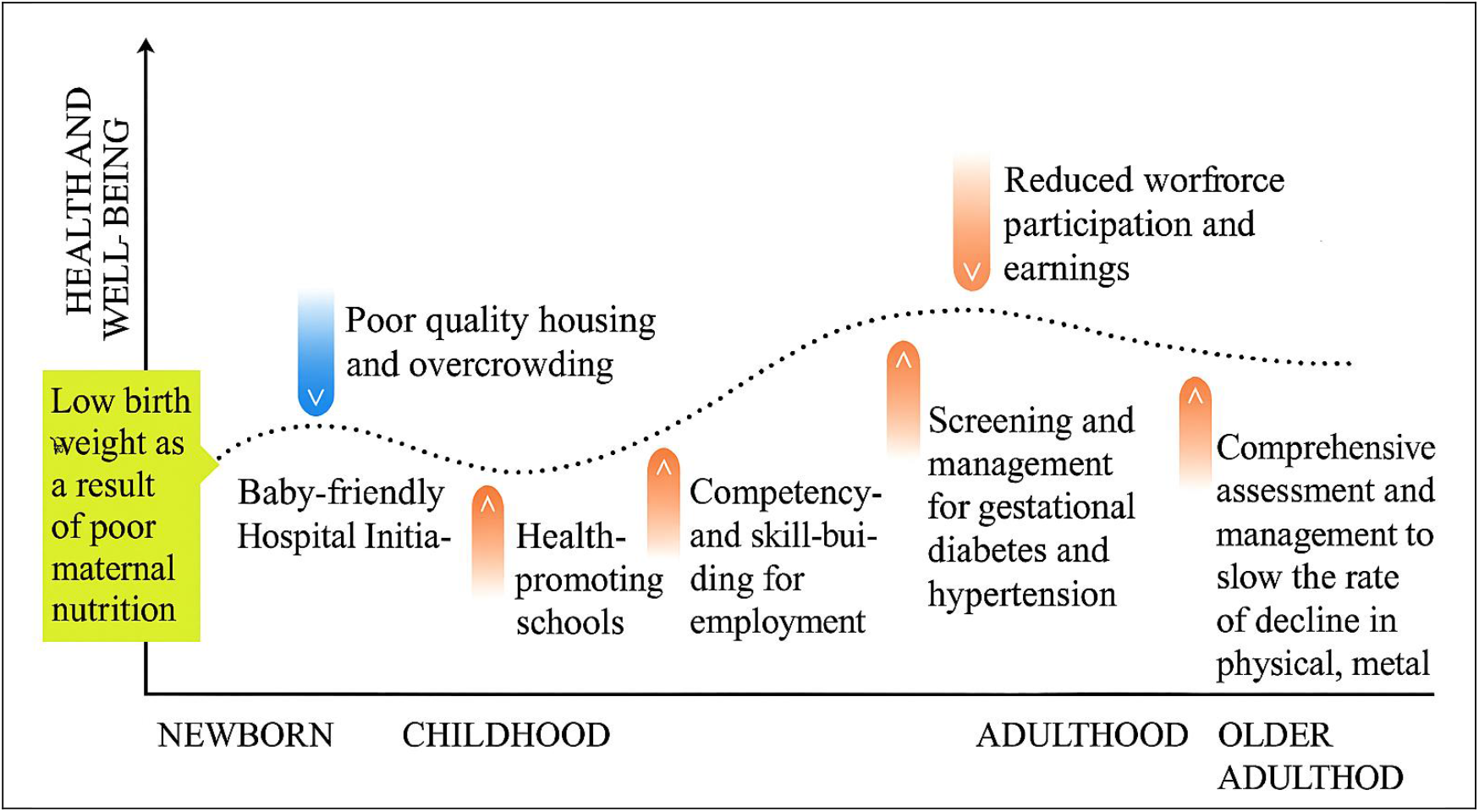
Two life-course frameworks: Lancet Commission & WHO (from (2))

Research over the past two decades has accumulated support for this framework.(2,7,11–16) Early studies conducted in high income countries demonstrated that childhood poverty is associated with poorer physical health, cognitive function and mental wellbeing in later-life.(7) A new generation of studies has expanded the geographies of analysis by combining studies in the family of U.S. Health and Retirement Studies. These cross-country investigations have yielded robust evidence that childhood poverty exerts a lasting influence on later-life health across a wide range of outcomes and contexts. In particular, analyses spanning nearly 30 countries have shown that adults whose childhood is marred by poverty go on to report higher probabilities of depression, worse episodic memory and increased risk of probable sarcopenia in later-life, even after accounting for in-between stages in life such as education achievement in early adulthood and later-life socioeconomic position or wealth.(12) Childhood poverty is not fully absorbed by adulthood conditions but retains a grasp on health outcomes decades later. Wahrendorf and Blane, for instance, studied CASP (Control, Autonomy, Self-realisation and Pleasure) in 13 countries in SHARE to conclude that the association of childhood disadvantage with later-life CASP remains.(17) This is despite education achievement and labour market positions in-between life stages. Notably, this study acknowledged the limitation arising from retrospective errors in childhood information, a vexing issue given its magnitude.

Frailty is another health outcome in focus.(13,18) A geriatric syndrome reflecting systemic physiological decline, across America, Britain and Europe, it is strongly associated with childhood poverty. Older adults who experienced poverty in childhood face significantly higher probabilities of frailty, with this association persisting across nearly 30 countries and remaining robust after accounting for social and health-related covariates. This syndrome is important because it presents a holistic measure of vulnerability in later-life.(13) Notably, this study accounted for the retrospective errors using latent construction, an emerging solution to the vexing issue.(7,11,15,16)

A broadening of the framework has been made recently, linking childhood poverty and later-life health by shifting to multidimensional conceptions of healthy ageing.(14) Drawing on six longitudinal ageing cohorts of 80,000 adults living in 32 countries it shows that childhood poverty is associated with reduced healthy ageing, as measured using latent trait models that integrate cognitive, physical, sensory and psychological functionings. The deleterious association persists even after adjusting for parental socioeconomic position, adult education attainment and labour market achievement, again stressing the imprint of childhood poverty.

Yet a deeper development of the framework goes ‘under the skin’, particularly with the concept of biological embedding.(19) Central to this development is the study of epigenetic processes, especially DNA methylation.(20,21) Evidence from the HRS venous blood sample indicates that childhood poverty is associated with accelerated epigenetic ageing.(12) The notion of epigenetic ageing offers a unifying framework for understanding the long and broad arm of childhood poverty. Rather than influencing a single outcome, childhood poverty appears to initiate systemic biological processes that manifest across domains and functions, demonstrating how social experiences are embodied then expressed in later-life outcomes.

Despite these advances, the literature remains characterised by a notable imbalance. The overwhelming majority of studies have focused on negative outcomes of disability, dysfunction, disease and clinical syndromes. While these outcomes are essential for understanding the burden of ill health, they offer a limited perspective on the ageing experience. Such a focus risks reinforcing a view of later-life as a period defined primarily by decline, overlooking the dimensions of life that contribute to positive wellbeing.(22,23)

In contrast, policy frameworks such as the WHO life-course framework emphasise a more holistic conception of ageing, centred on functional ability and quality of life.(10) This emphasis draws attention to positive dimensions of ageing viz. what older adults can do and experience, not merely what they lack. Quality of life assumes particular importance here. The CASP instrument which taps into dimensions of control, autonomy, self-realisation and pleasure is a validated measure of quality of life or positive wellbeing that is specific to older populations.(22) It captures dimensions central to human flourishing – ability to exert control over circumstances, maintain independence, realise personal potential and derive enjoyment from life.

This study aims to answer these questions. Does childhood poverty shape old age quality of life as measured using CASP, accounting for adult socioeconomic positions? How does this shaping vary across 29 countries with different welfare regimes and health systems? It rises to the challenge above viz. the achievements of ageing societies go beyond extending life to enhancing its quality: lives with control, autonomy, self-realisation and pleasure.

## MATERIALS AND METHODS

This retrospective and prospective study draws on a family of longitudinal ageing studies: the U.S. Health and Retirement Study (HRS wave 11, 2016), the English Longitudinal Study of Ageing (ELSA waves 8 and 11, 2016 and 2025), the Survey of Health, Ageing and Retirement in Europe (SHARE wave 6, 2015), and The Irish Longitudinal Study on Ageing (TILDA wave 1, 2009– 2011).(24–27) Together these surveyed 59,569 older adults (over 50) in 29 countries, spanning varied histories, welfare regimes and health systems. The design follows recent research on cross-national life-course shaping of health in which retrospectively recalled childhood conditions are linked with prospectively measured outcomes in later-life.(2,12–14)

The outcome is quality of life measured using CASP (control, autonomy, self-realisation and pleasure), four domains elicited with twelve items to capture positive wellbeing in later-life rather than disease or dysfunction.(22) (The 19 items version is not uniformly available; moreover, the HRS used a slightly different instrument of control, autonomy, self-expression and mastery (CASM), rescaled to have the same range as CASP).(24) Two notes on the use of two instruments (CASP, CASM). First, although they are slightly different, this approach is increasingly common. For instance, Courtin and colleagues have used CESD and Euro-D (slightly different) for depression, Tampubolon and Maharani have used different biomarkers for allostatic load, and Tampubolon have used two different sets of items for healthy ageing.(2,28–30) The rationale rests on the focus on risk factor association rather than on prevalence. Second, sensitivity analysis, separating HRS, is carried out to assess the robustness of the combined ageing studies. Following earlier work using CASP trajectory in Britain, the scale is oriented so that higher scores indicate more control, autonomy, self-realisation and pleasure.(23)

The key exposure is childhood poverty which has been fruitfully constructed for life-course research.(2,7,11–14) Participants recalled childhood material circumstances, typically around ages 10 to 14. Indicators differ across surveys but include number of books, rooms and people in the childhood home, overcrowding, housing facilities, access to running water or heating, and financial hardship or family indebtedness. These retrospective reports are not assumed to be error-free. Consistent with recent research on the life-course shaping of depression, episodic memory, probable sarcopenia, frailty, multimorbidity and healthy ageing, childhood poverty is entered as a binary latent class (poor or advantaged).(2,7,12–14) This approach treats each recalled childhood indicator as an imperfect signal of underlying material deprivation and permits comparison despite differences in indicators; please see the most recent details here.(2) The covariate set is the same as those used in recent literature on the life-course shaping of later-life health. Models adjust for age, sex, education, marital status, current wealth and country. These covariates represent adult and later-life stages and help distinguish the association of childhood poverty from pathways through education, adult socioeconomic achievement and current wealth.

## Statistical analysis

A base model (age, sex, childhood poverty, country) and a complete model were fitted, focusing on the attenuation of poverty coefficient across the two models. This has proven an effective strategy in studying childhood poverty’s association with later-life health.(7,17) The models estimate the association between latent childhood poverty and CASP using country fixed effects linear model.(2,12–14) Country coefficients absorb unobserved national differences in histories, health systems and welfare regimes. Because childhood poverty is an estimated latent exposure, standard errors are corrected.(31,32) Then, sensitivity analyses compare fixed- and random-effects linear models, stratify by sex, and examine each cohort study separately. All estimation is done in Latent GOLD Syntax 6.1 and significance is set at 5%.(33)

## RESULTS

Table 1 presents the summary statistics for the analytic sample comprising 59,569 older adults living in communities in 29 high-income countries. The analytic sample has large proportions from England (11.2%), Ireland (5.8%) and the U.S. (5.6%). The sex distribution consists of 33,976 women (57.0%) and 25,593 men (43.0%); average age is 66.4 years (SD = 9.5), thus within later-life stage.

**Table 1.** Descriptive summaries of analytic sample.

|  | Male | Female | Total |
|---|---|---|---|
|  | N=25593 | N=33976 | N=59569 |
| Childhood poverty |  |  |  |
| Childhood advantaged | 19873 (77.7%) | 26042 (76.6%) | 45915 (77.1%) |
| Childhood poor | 5720 (22.3%) | 7934 (23.4%) | 13654 (22.9%) |
| CASP-12 | 38.1 (6.1) | 37.5 (6.4) | 37.7 (6.3) |
| Age | 66.6 (9.2) | 66.3 (9.7) | 66.4 (9.5) |
| Education |  |  |  |
| Up to Primary | 7899 (30.9%) | 12479 (36.7%) | 20378 (34.2%) |
| High school | 10614 (41.5%) | 13492 (39.7%) | 24106 (40.5%) |
| College/more | 7080 (27.7%) | 8005 (23.6%) | 15085 (25.3%) |
| Marital status |  |  |  |
| Single | 1540 (6.0%) | 1689 (5.0%) | 3229 (5.4%) |
| Married/Partnered | 19072 (74.5%) | 20208 (59.5%) | 39280 (65.9%) |
| Sep/Widowed | 4981 (19.5%) | 12079 (35.6%) | 17060 (28.6%) |
| Wealth quartile |  |  |  |
| Above bottom quartile | 22100 (86.4%) | 27460 (80.8%) | 49560 (83.2%) |
| Bottom quartile | 3493 (13.6%) | 6516 (19.2%) | 10009 (16.8%) |
| country |  |  |  |
| Ireland | 1572 (6.1%) | 1876 (5.5%) | 3448 (5.8%) |
| England | 2909 (11.4%) | 3743 (11.0%) | 6652 (11.2%) |
| Austria | 700 (2.7%) | 1073 (3.2%) | 1773 (3.0%) |
| Germany | 1584 (6.2%) | 1767 (5.2%) | 3351 (5.6%) |
| Sweden | 1202 (4.7%) | 1404 (4.1%) | 2606 (4.4%) |
| Netherlands | 612 (2.4%) | 802 (2.4%) | 1414 (2.4%) |
| Spain | 1150 (4.5%) | 1418 (4.2%) | 2568 (4.3%) |
| Italy | 1370 (5.4%) | 1702 (5.0%) | 3072 (5.2%) |
| France | 1208 (4.7%) | 1634 (4.8%) | 2842 (4.8%) |
| Denmark | 1246 (4.9%) | 1455 (4.3%) | 2701 (4.5%) |
| Greece | 321 (1.3%) | 421 (1.2%) | 742 (1.2%) |
| Switzerland | 1001 (3.9%) | 1216 (3.6%) | 2217 (3.7%) |
| Belgium | 1442 (5.6%) | 1736 (5.1%) | 3178 (5.3%) |
| Israel | 277 (1.1%) | 402 (1.2%) | 679 (1.1%) |
| Czechia | 1144 (4.5%) | 1771 (5.2%) | 2915 (4.9%) |
| Poland | 1204 (4.7%) | 1526 (4.5%) | 2730 (4.6%) |
| Luxembourg | 342 (1.3%) | 427 (1.3%) | 769 (1.3%) |
| Hungary | 251 (1.0%) | 393 (1.2%) | 644 (1.1%) |
| Slovenia | 867 (3.4%) | 1304 (3.8%) | 2171 (3.6%) |
| Estonia | 924 (3.6%) | 1721 (5.1%) | 2645 (4.4%) |
| Croatia | 472 (1.8%) | 619 (1.8%) | 1091 (1.8%) |
| Lithuania | 456 (1.8%) | 830 (2.4%) | 1286 (2.2%) |
| Bulgaria | 340 (1.3%) | 517 (1.5%) | 857 (1.4%) |
| Cyprus | 168 (0.7%) | 287 (0.8%) | 455 (0.8%) |
| Finland | 494 (1.9%) | 570 (1.7%) | 1064 (1.8%) |
| Latvia | 261 (1.0%) | 449 (1.3%) | 710 (1.2%) |
| Malta | 324 (1.3%) | 412 (1.2%) | 736 (1.2%) |
| Slovakia | 430 (1.7%) | 505 (1.5%) | 935 (1.6%) |
| USA | 1322 (5.2%) | 1996 (5.9%) | 3318 (5.6%) |

Childhood poverty is prevalent: one in five (22.9%) grew up in poverty, higher among women (23.4%) than men (22.3%). The outcome CASP shows an overall mean of 37.7 (SD = 6.3) out of a maximum of 48 – more frequent than ‘sometimes’ across all four dimensions. On adult education levels, two-in-five completed high school, and one-in-four finished college; a substantial majority are married or partnered (65.9%), while 16.8% currently in the bottom wealth quartile.

### Base and complete models

Table 2 presents models predicting CASP scores, using both fixed-effects and random-effects/intercepts models. To interpret the direct impact of childhood poverty the models are presented in two stages: base model (adjusting only for childhood poverty, age, sex and country) and complete model (adding adult education, marital status and current wealth). The attenuation in childhood poverty coefficient is clear. The consistency between the fixed and random effects models is also striking, stressing robust associations. The fixed effect model is thus preferred and interpreted.

**Table 2.** Base and complete models, each with fixed and random effects.

|  | Base | Complete | Base | Complete |
|---|---|---|---|---|
|  | Fixed | Fixed effects | Random | Random effects |
|---|---|---|---|---|
| Childhood poor | -1.168***<br>(-19.04) | -0.685***<br>(-11.15) | -1.173***<br>(-19.12) | -0.689***<br>(-11.21) |
| Sex, Female | -0.534***<br>(-11.10) | -0.254***<br>(-5.26) | -0.535***<br>(-11.11) | -0.254***<br>(-5.27) |
| Age | -0.095***<br>(-36.36) | -0.074***<br>(-27.87) | -0.094***<br>(-36.35) | -0.074***<br>(-27.86) |
| High school |  | 1.150***<br>(19.12) |  | 1.151***<br>(19.13) |
| College/more |  | 2.028***<br>(29.78) |  | 2.028***<br>(29.79) |
| Married/Partnered |  | 0.950***<br>(8.98) |  | 0.947***<br>(8.96) |
| Sep/Widowed |  | 0.368***<br>(3.29) |  | 0.367***<br>(3.28) |
| Bottom quartile |  | -1.926***<br>(-25.72) |  | -1.935***<br>(-25.86) |
| Constant | 44.81***<br>(234.90) | 41.99***<br>(189.79) | 44.14***<br>(104.10) | 41.13***<br>(96.90) |
| Country dummies | Yes | Yes | No | No |
| Observations | 59569 | 59569 | 59569 | 59569 |
| Adjusted *R*<sup>2</sup> | 0.143 | 0.17 | 0.09 | 0.10 |

The base model shows that childhood poverty exerts a severe penalty on later-life positive wellbeing, reduced CASP by 1.2 points (p < 0.01). When adult and later-life socioeconomic positions are introduced (complete model), the penalty attenuates but remains highly significant, a 0.7 points reduction (p < 0.01). This attenuation indicates that while educational and financial achievements in adulthood and later-life partially account for the association of childhood poverty, they do not erase it. Childhood poverty maintains a direct, independent, and lasting association with reduced control, autonomy, self-realisation, and pleasure in later-life, providing strong evidence for the life-course shaping of positive wellbeing. The biological embedding of early material hardship appears to leave an indelible mark on psychosocial wellbeing many decades later.

Beyond childhood conditions, the complete model reveals the powerful influence of adulthood social determinants. Educational attainment serves as a considerable protection; compared to those with only primary education, completing high school boosts CASP by one point, while finishing college provides a two-point advantage. These educational premium offsets the childhood poverty penalty, promising education as a critical lever for life-course wellbeing. Similarly, being married or partnered confers a strong psychosocial benefit (0.9 points) compared to being single.

Current financial disadvantage translates to wellbeing. Being in the bottom wealth quartile in later-life associates with 1.9-point lower CASP, almost as large as the college premium. Demographic penalties are also evident: women score significantly lower than men (0.25 points in the complete model), and advancing age steadily erodes quality of life by 0.07 points per year. The move from base to complete model improves the explanatory power (R-squared) from 0.14 to 0.17, consistent with later-life quality of life being an accumulation of life-course exposures rather than the product of any single life stage.

### Cross-national age profiles and the Nordic model

Figure 2 shows the predicted CASP profiles for adults spanning the ages of 70 to 90 across the 29 high-income nations, distinguishing the childhood poor (dashed lines) from the childhood advantaged (solid lines). The trellis plot captures the ubiquity of the life-course shaping of health – childhood poverty matters. In every country the dashed line remains below the solid line. This gap expresses the childhood poverty penalty, transcending highly varied histories, healthcare systems and welfare regimes. Also, every country presents a downward slope, reinforcing the finding that advancing age everywhere erodes positive wellbeing.

**Figure 2.**
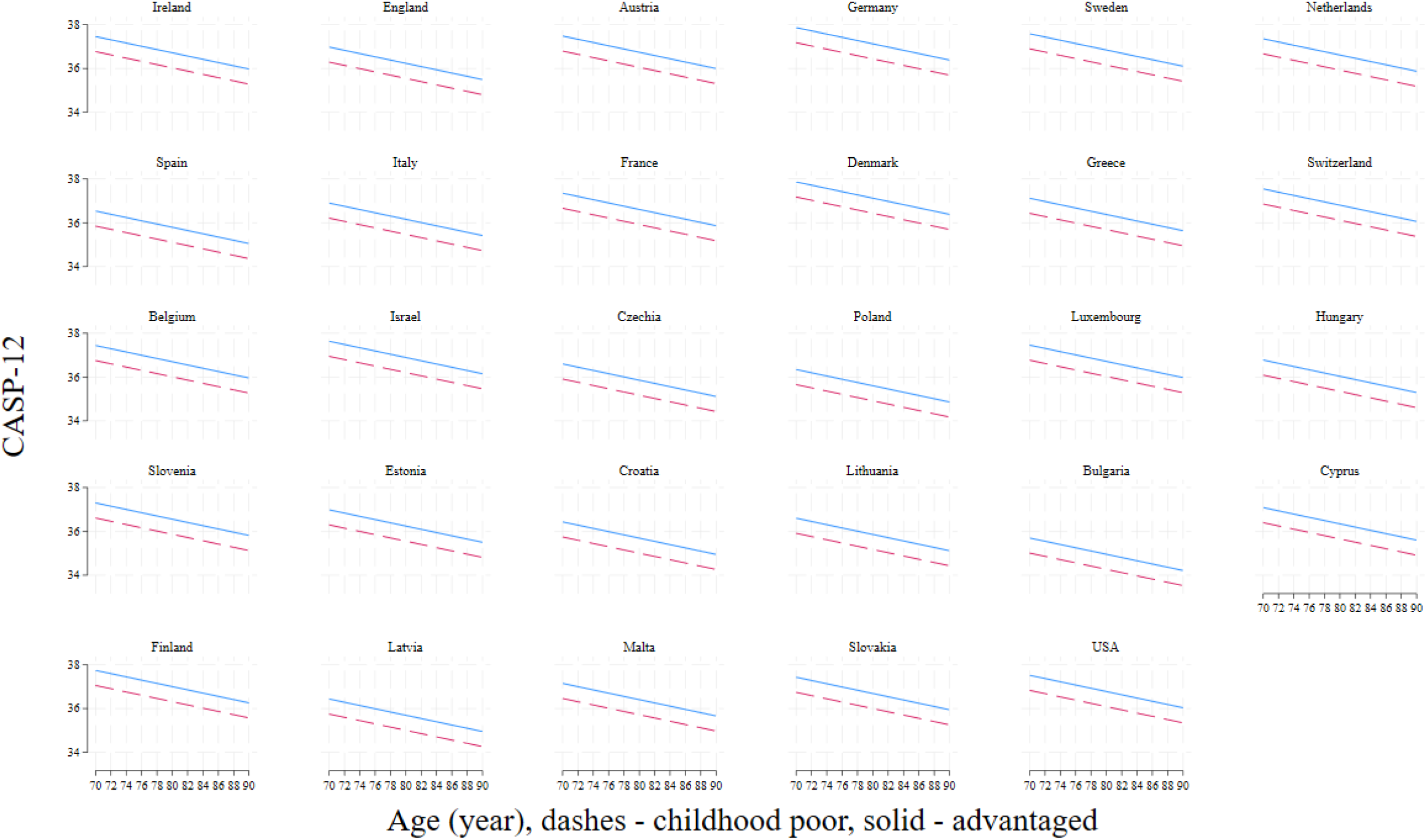
Marginal predictions of CASP (control, autonomy, self-realisation, pleasure) for ages 70–90 across 29 nations, dashed = childhood poor; solid = childhood advantaged.

Strikingly, the levels of CASP differ markedly across nations: the Nordic advantage is evident. In these Nordic nations, the intercept for the entire population is elevated. Focus on the bottom left: countries such as Finland have visibly higher CASP scores at age 70 compared to Latvia (Eastern Europe). In fact, Finns aged 90 fare equally with Latvians two decades younger. I have discovered this to be the case also in other negative health outcomes.(12–14) But importantly, while these robust welfare states elevate the levels for all citizens, they do not completely close the parallel gap started by childhood poverty. The life-course imprint remains visible even within the most effective institutional contexts, repeating the call to eradicate child poverty globally.

In the supplement I present sensitivity analyses including stratification by sex and by cohort study. Notably, the main results remain.

### DISCUSSION AND CONCLUSION

On both sides of the Atlantic I found the childhood poor had less control, autonomy, self-realization and pleasure in later-life. Because CASP captures positive wellbeing rather than absence of illness, the finding broadens the reach of the life-course framework underpinning the WHO Decade of Healthy Ageing.(4) It extends recent evidence on depression, episodic memory, probable sarcopenia, frailty, epigenetic age, multimorbidity and healthy ageing,(2,7,12–14) and is achieved by faithfully using childhood overcrowding indicators elaborated in the WHO Framework above.(10) The framework guides this contribution in another way. Later-life CASP is located within a sequence of exposures beginning in childhood and extending through adult education and labour market achievement as well as later-life wealth. The attenuation of childhood poverty coefficient is therefore informative: adult conditions account for the association but do not completely remove the childhood poverty penalty.

The association is observed across diverse settings (the U.K.’s National Health Service to the U.S.’ primarily private health insurance, and everything in between). This contrast shows that the childhood poverty penalty in later-life CASP is not reducible to one health system. The CASP marginal plots show both ubiquitous childhood poverty penalty and important national variations. In particular, the Nordic advantage suggests that welfare institutions and social protection may critically influence the level of later-life positive wellbeing, enriching the manifestation of the Nordic welfare model.(9,15)

Following recent research, the study implements a latent construct to account for error in retrospective childhood information.(7,15,16,34) Recent analyses discovered large errors in retrospective report of childhood overcrowding emphasised in the WHO Framework:(10) two thirds of Britons aged 50 incorrectly recalled overcrowding when they were eleven.(7,14,35) Therefore, treating childhood poverty as a latent construct built from imperfect indicators is preferable to treating childhood indicators like any other error-free covariates. Retrospective information should not be discarded, though, because it remains indispensable for life-course research. But use them with methodological discipline. The latent construct approach enables cross-country designs when childhood indicators differ, while country controls absorb national welfare regimes and health systems.

This accounting for measurement error is one advance over past research on CASP.(17,23) Other advances include more sister studies (four), more countries (29)and more recent data. All points to childhood poverty goes with lower CASP in later-life. In the same issue with Wahrendorf and Blane,(17) I derived trajectories of CASP from 2002 to 2010 (23) – and this points to further research. Beyond association with lower CASP scores, does childhood poverty quicken losses in control, autonomy, self-realisation or pleasure around the world?

Past evidence has been inconclusive – we have found on inflammation trajectories this not to be the case(36) while others have found on frailty trajectories this to be the case.(18) The rate of losses in control, autonomy, self-realisation and pleasure among the childhood poor is an eminently viable research focus. This study has also revealed striking cross-country differences in CASP scores, equivalent to two-decades CASP scores difference at the same calendar age. This calls for further investigation as done for mortality and ill health.(37)

### Strengths and weaknesses

This study builds on many strengths including harmonised sister studies, a large analytic sample, 29-country coverage, a positive CASP outcome, latent construct of childhood poverty, and its indicators exemplified in the WHO Framework.(10) However, it has weaknesses including observational design which prevent causal interpretation of the coefficient, healthy survivor bias attenuating the association, imperfect harmonisation of outcome (HRS instead collected measures of control, autonomy, self-expression and mastery), lack of psychological childhood adversity, and the absence of a structural/path-analytic model. Though these caution against causal statement, these do not undermine the consistency of the childhood poverty association across the largest number of countries to date.

### Health policy for a more pleasurable later-life?

Positive wellbeing in later-life should be welcomed as a life-course achievement. Thus, policies to improve ageing experience cannot start at retirement or even midlife, but must include child poverty reduction and secure housing as a baseline healthy ageing intervention.(2,7) The study therefore supports the WHO Decade of Healthy Ageing by showing that functional ability and positive wellbeing in later-life are shaped by exposures long before it starts.(4)

The cross-national evidence uniquely suggests that health systems alone are insufficient. The contrast between the U.S. and U.K. health systems illustrates that health-care financing is only one part of the institutional environment shaping later-life CASP.(38) Wider welfare arrangements, income protection, education systems and housing policies may be as important. The Nordic advantage in predicted CASP profiles is consistent with the evidence that societies with stronger social protection and more universalist institutions may support higher wellbeing for older adults, including those who grew up poor decades earlier.(14,15,39)

Policy research on ageing increasingly relies on retrospective decades-long life history interviews; they are indispensable for understanding the life-course shaping of health.(7,17,18) But the magnitude of recall error means that evidence should be produced using measurement strategies that acknowledge this.(2,7,14,35) The policy message is twofold: reducing child poverty is a long-term investment for later-life wellbeing, and robust measurement is essential for reaping the fruit of such investment.

The study complements a recent work on the life-course shaping of healthy ageing in an expanded set of countries (including middle income countries).(2,14) The policy take then becomes stronger. Childhood poverty matters both for whether older adults retain functionings,(2) and whether they experience later-life with control, autonomy, self-realisation and pleasure. The lifelong reward of eliminating child poverty is not only fewer illnesses or even better functionings, but a flourishing later-life.

## Data Availability

All data produced are available online at https://hrs.isr.umich.edu (HRS), www.share-project.org (SHARE), www.elsa-project.ac.uk (ELSA), https://tilda.tcd.ie (TILDA).

https://www.share-project.org

https://www.elsa-project.ac.uk

https://tilda.tcd.ie

## BIBLIOGRAPHY

1. United Nations. World Population Prospects [Internet]. 2022 [cited 2022 Jun 27]. Available from: https://population.un.org/wpp/Download/SpecialAggregates/Geographical/

2. Tampubolon G. Parental socioeconomic position, childhood poverty and healthy ageing: retrospective and prospective study of six cohort surveys across 32 wealthy and developing nations. Front Public Health. 2026 Jul 17. doi:10.3389/fpubh.2026.1879570

3. WHO. World Report on Ageing and Health. Geneva: WHO; 2015.

4. WHO. UN Decade of Healthy Ageing 2021–2030 [Internet]. 2021 [cited 2021 Jan 14]. Available from: https://www.who.int/initiatives/decade-of-healthy-ageing

5. Elder GH. The life course as developmental theory. Child Dev. 1998 Feb;69(1):1–12.

6. Dannefer D. Cumulative advantage/disadvantage and the life course: cross-fertilizing age and social science theory. J Gerontol Ser B. 2003 Nov 1;58(6):S327–37. doi:10.1093/geronb/58.6.S327

7. Tampubolon G. Growing up in poverty, growing old in infirmity: The long arm of childhood conditions in Great Britain. PLoS One. 2015;10(12):e0144722. doi:10.1371/journal.pone.0144722

8. Kuh D, Ben-Shlomo Y, Lynch J, Hallqvist J, Power C. Life course epidemiology. J Epidemiol Community Health. 2003 Oct 1;57(10):778–83. doi:10.1136/jech.57.10.778 PubMed PMID: 14573579.

9. Siegrist J, Marmot M, editors. Social Inequalities in Health: New Evidence and Policy Implications. Oxford: Oxford University Press, U.S.A.; 2010. 270 p.

10. WHO. Framework to Implement a Life Course Approach in Practice [Internet]. [cited 2025 Jul 9]. Available from: https://www.who.int/publications/i/item/9789240112575

11. Tampubolon G. Recall error and recall bias in life course epidemiology [MPRA Paper] [Internet]. 2010 [cited 2022 Feb 2]. Available from: https://mpra.ub.uni-muenchen.de/23847/

12. Tampubolon G. The long arm hypothesis: childhood poverty, epigenetic ageing, and late-life health in America, Britain, and Europe. In: Hoffmann R, editor. Handbook of Health Inequalities across the Life Course. Cheltenham: Edward Elgar; 2023. p. 252–74. doi:10.4337/9781800888166.00026

13. Tampubolon G. Growing up in poverty, growing old in frailty: the life course shaping of health in the United States, England and Europe—a prospective and retrospective study. Sci Rep. 2025 May 3;15(1):15510. doi:10.1038/s41598-025-99929-2

14. Tampubolon G. Growing up in poverty, growing old with multimorbidity in America, Britain, China, Europe and Indonesia: a retrospective and prospective study. In: Asiamah N, Khan H, Nesser W, Alvarez EN, editors. Morbidity Prevention in Ageing [Internet]. New York, NY: Elsevier; 2026 [cited 2025 Apr 7]. p. Chapter 16. Available from: https://www.medrxiv.org/content/10.1101/2024.07.16.24310497v1

15. Lennartsson C, Eyjólfsdóttir HS, Celeste RK, Fritzell J. Social class and infirmity. The role of social class over the life-course. SSM - Popul Health. 2018 Apr 1;4:169–77. doi:10.1016/j.ssmph.2017.12.001

16. Pakpahan E, Hoffmann R, Kröger H. The long arm of childhood circumstances on health in old age: Evidence from SHARELIFE. Adv Life Course Res. 2017 Mar 1;31:1–10. doi:10.1016/j.alcr.2016.10.003

17. Wahrendorf M, Blane D. Does labour market disadvantage help to explain why childhood circumstances are related to quality of life at older ages? Results from SHARE. Aging Ment Health. 2015 Jul 3;19(7):584–94. doi:10.1080/13607863.2014.938604 PubMed PMID: 25033373.

18. Van Der Linden BWA, Sieber S, Cheval B, Orsholits D, Guessous I, Gabriel R, et al. Life-course circumstances and frailty in old age within different European welfare regimes: A longitudinal study with SHARE. J Gerontol Ser B. 2020 Jul 1;75(6):1326–35. doi:10.1093/geronb/gbz140

19. Hertzman C. The biological embedding of early experience and its effects on health in adulthood. Ann N Y Acad Sci. 1999;896:85–95. doi:10.1111/j.1749-6632.1999.tb08107.x PubMed PMID: 10681890.

20. Widagdo J, Anggono V. The m6A-epitranscriptomic signature in neurobiology: from neurodevelopment to brain plasticity. J Neurochem. 2018;147(2):137–52. doi:10.1111/jnc.14481

21. Szyf M. How do environments talk to genes? Nat Neurosci. 2013 Jan;16(1):1. doi:10.1038/nn.3286

22. Hyde M, Higgs P, Wiggins RD, Blane D. A decade of research using the CASP scale: key findings and future directions. Aging Ment Health. 2015 Jul 3;19(7):571–5. doi:10.1080/13607863.2015.1018868 PubMed PMID: 25847497.

23. Tampubolon G. Delineating the third age: joint models of older people’s quality of life and attrition in Britain 2002-2010. Aging Ment Health. 2015 Jul;19(7):576–83. doi:10.1080/13607863.2014.1003279.

24. Sonnega A, Faul JD, Ofstedal MB, Langa KM, Phillips JW, Weir DR. Cohort Profile: the Health and Retirement Study (HRS). Int J Epidemiol. 2014 Apr 1;43(2):576–85. doi:10.1093/ije/dyu067

25. Donoghue OA, McGarrigle CA, Foley M, Fagan A, Meaney J, Kenny RA. Cohort Profile Update: The Irish Longitudinal Study on Ageing (TILDA). Int J Epidemiol. 2018 Oct 1;47(5):1398–1398l. doi:10.1093/ije/dyy163

26. Steptoe A, Breeze E, Banks J, Nazroo J. Cohort Profile: The English Longitudinal Study of Ageing. Int J Epidemiol. 2013 Dec 1;42(6):1640–8. doi:10.1093/ije/dys168

27. Börsch-Supan M, Börsch-Supan A, Andersen-Ranberg K, Borbye-Lorenzen N, Cofferen J, Deza-Lougovski YI, et al. Cohort Profile Update: Survey of Health, Ageing and Retirement in Europe –Biomarker data for age-related health conditions [Internet]. Munich: Munich Research Institute for the Economics of Aging and SHARE Analyses (MEA); 2026 Jan [cited 2026 May 12]. p. 2026.01.28.26344911. Available from: https://www.medrxiv.org/content/10.64898/2026.01.28.26344911v1 doi:10.64898/2026.01.28.26344911

28. Courtin E, Knapp M, Grundy E, Avendano-Pabon M. Are different measures of depressive symptoms in old age comparable? An analysis of the CES-D and Euro-D scales in 13 countries. Int J Methods Psychiatr Res. 2015 Dec 2;24(4). doi:10.1002/mpr.1489

29. Tampubolon G, Maharani A. Trajectories of allostatic load among older Americans and Britons: longitudinal cohort studies. BMC Geriatr. 2018 Oct 23;18(1):255. doi:10.1186/s12877-018-0947-4

30. Li G, Rubeis VD, Cesari M, Sadana R, Jacob CM, Lee H, et al. Life course shaping of allostatic load across 17 countries: Evidence from England, the U.S., Europe, China and Indonesia. medRxiv; 2026. doi:10.64898/2026.07.29.26359214

31. Vermunt JK. Latent class modeling with covariates: Two improved three-step approaches. Polit Anal. 2010 Jan 4;18(4):450–69. doi:10.1093/pan/mpq025

32. Vermunt JK. Stepwise estimation of latent variable models: An overview of approaches. Stat Model. 2025 Dec 1;25(6):530–51. doi:10.1177/1471082X251355693

33. Vermunt JK, Magidson J. LG-Syntax User’s Guide version 6.1. Tilburg, Netherland: Statistical Innovations Europe; 2025.

34. Berndt H, Fors S. Childhood living conditions, education and health among the oldest old in Sweden. Ageing Soc. 2016 Mar;36(3):631–48. doi:10.1017/S0144686X14001470

35. Tampubolon G. Social mobility and long-term episodic memory in Britain [Internet]. medRxiv; 2026 [cited 2026 Jun 18]. p. 2026.04.12.26350709. Available from: https://www.medrxiv.org/content/10.64898/2026.04.12.26350709v1 doi:10.64898/2026.04.12.26350709

36. Tampubolon G, Fajarini M. Does a poor childhood associate with higher and steeper inflammation trajectories in the English Longitudinal Study of Ageing? [Internet]. bioRxiv; 2018 [cited 2018 Feb 19]. p. 267559. Available from: https://www.biorxiv.org/content/10.1101/267559v1 doi:10.1101/267559

37. Mackenbach JP. Socio-economic inequalities in health in western Europe: From description to explanation to intervention. In: Siegrist J, Marmot M, editors. Social Inequalities in Health: New evidence and policy implications. Oxford University Press; 2006. p. 223–50.

38. Britnell M. In Search of the Perfect Health System. London: Palgrave; 2015.

39. Maharani A, Dawes P, Nazroo J, Tampubolon G, Pendleton N, the SENSE-Cog WP1 group. Healthcare system performance and socioeconomic inequalities in hearing and visual impairments in 17 European countries. Eur J Public Health. 2021 Feb 1;31(1):79–86. doi:10.1093/eurpub/ckaa155

