## Supplement for "The life course shaping of quality of life in older age: does childhood poverty associate with control, autonomy, self-realisation and pleasure? A retrospective and prospective study of HRS, ELSA, SHARE and TILDA cohorts"

This supplement presents additional analyses supporting the main findings reported in the main manuscript. The analyses below assess whether the main negative association between childhood poverty and CASP is robust across sex and across the four cohort studies. In sex-stratified fixed-effects models, childhood poverty remains negatively associated with CASP among both women and men. In cohort-stratified models, the childhood poverty coefficient is negative in the HRS, ELSA, SHARE and TILDA samples, with the strongest estimate observed in SHARE and statistically significant associations in all four cohorts. Together, these analyses indicate that the life-course association between childhood poverty and reduced later-life control, autonomy, self-realisation and pleasure is not driven by one sex or by a single ageing study, strengthening the interpretation that the main results reflect a broad and reproducible pattern across high-income countries.

Table S1. Fixed effects linear models explaining CASP (Control, Autonomy, Self-realisation and Pleasure), stratified by sex

| | Female | Male |
|---|---|---|
| Childhood poor | -0.779*** | -0.566*** |
| | (-9.46) | (-6.14) |
| Age | -0.0792*** | -0.0687*** |
| | (-22.21) | (-17.00) |
| High school | 1.124*** | 1.129*** |
| | (14.11) | (12.24) |
| College/more | 2.027*** | 1.969*** |
| | (21.82) | (19.45) |
| Married/Partnered | 0.715*** | 1.184*** |
| | (4.82) | (7.86) |
| Sep/Widowed | 0.105 | 0.706*** |
| | (0.69) | (4.19) |
| Bottom quartile | -1.806*** | -2.152*** |
| | (-18.76) | (-17.94) |
| Constant | 42.55*** | 41.11*** |
| | (142.27) | (127.37) |
| Observations | 33976 | 25593 |
| Adjusted $R^2$ | 0.187 | 0.152 |

*t* statistics in parentheses

* p<0.1, ** p<0.05, *** p<0.01

Table S2. Fixed effects linear model explaining CASP( Control, Autonomy, Self-realisation and Pleasure), stratified by cohort study

| | US/HRS | England/ ELSA | Europe/ SHARE | Ireland/ TILDA |
|---|---|---|---|---|
| Childhood poor | -0.824* | -0.588*** | -1.230*** | -0.681*** |
| | (-1.86) | (-3.12) | (-18.51) | (-3.09) |
| Sex, Female | -0.340 | 0.315** | -0.388*** | 0.498*** |
| | (-1.22) | (2.23) | (-6.87) | (2.85) |
| Age | -0.088*** | -0.042*** | -0.075*** | 0.057*** |
| | (-5.91) | (-6.30) | (-23.98) | (5.27) |
| High school | 1.012* | 0.463*** | 1.599*** | 0.873*** |
| | (1.83) | (2.67) | (25.05) | (3.87) |
| College/more | 2.426*** | 1.371*** | 2.510*** | 1.883*** |
| | (4.51) | (7.87) | (33.19) | (6.67) |
| Married/Partnered | 0.560 | 1.252*** | 0.538*** | 1.002*** |
| | (0.31) | (4.04) | (4.27) | (3.37) |
| Sep/Widowed | 0.484 | 0.609* | 0.181 | 0.192 |
| | (0.88) | (1.91) | (1.34) | (0.55) |
| Bottom quartile | -3.303*** | -2.897*** | -2.956*** | -1.145*** |
| | (-6.95) | (-16.80) | (-38.71) | (-3.55) |
| Constant | 42.57*** | 39.82*** | 41.87*** | 35.06*** |
| | (33.53) | (69.66) | (169.05) | (44.55) |
| Observations | 3318 | 6652 | 46151 | 3448 |
| Adjusted $R^2$ | 0.041 | 0.076 | 0.118 | 0.035 |

*t* statistics in parentheses

* p<0.1, ** p<0.05, *** p<0.01
